# Public perceptions of non-adherence to COVID-19 measures by self and others in the United Kingdom

**DOI:** 10.1101/2020.11.17.20233486

**Authors:** Simon N Williams, Christopher J. Armitage, Tova Tampe, Kimberly Dienes

**Affiliations:** Centre for People and Organisation, School of Management, Swansea University, Swansea, SA1 8EN; Department of Medical Social Sciences, Feinberg School of Medicine, Northwestern University, Chicago, Illinois, 60611, United States of America; Manchester Centre for Health Psychology, University of Manchester, Manchester, M13 9PL; Manchester University NHS Foundation Trust, Manchester Academic Health Science Centre, Manchester, M13 9PL; World Health Organisation, 1211 Geneva 27, Switzerland; Department of Psychology, School of Human and Health Sciences, Swansea University, Swansea, SA2 8PP; Honorary Lecturer, Manchester Centre for Health Psychology, Division of Psychology and Mental Health, University of Manchester

## Abstract

**OBJECTIVE:** To explore the perceptions of non-adherence to COVID-19 policy measures by self and others in the UK, focusing on perceived reasons for non-adherence.

**DESIGN:** Qualitative study comprising 12 online focus groups conducted between 25th September and 13th November 2020.

**SETTING:** Online video-conferencing

**PARTICIPANTS:** 51 UK residents aged 18 and above, reflecting a range of ages, genders and race/ethnicities.

**RESULTS:** Participants reported seeing an increase in non-adherence in others and identified a number of challenges to their own adherence to measures. Thematic analysis identified six main themes related to participants’ reported reasons for non-adherence in self and others: (1) Alert fatigue (2) Inconsistent rules (3) Lack of trust in government (4) Helplessness (5) Resistance and rebelliousness (6) Reduced perception of risk and the prospect of a vaccine. Participants also raised concerns that adherence would be impacted by a desire to socialise over Christmas. Two forms of non-adherence were observed: overt rule-breaking and subjective rule interpretation.

**CONCLUSIONS:** Adherence may be improved by: less frequent and clearer information on COVID-19 to reduce alert fatigue; implementing a more unified set of measures within and across countries in the UK; role modelling good adherence by authority figures; exploring ways to mitigate the impact that events like Christmas vaccine ‘breakthroughs’ may have on reducing adherence.

## BACKGROUND

The current coronavirus (COVID-19) pandemic continues to present a significant challenge to global public health. As of November 2020, no vaccination for the virus has been approved for use, and as such, non-pharmaceutical, social distancing measures provide the best means through which to control the transmission of the virus and its associated morbidity and mortality. Social distancing measures used in the UK include but are not limited to: keeping physically separate (1-2 meters in the UK), only meeting with others in ways permitted under current legislation, and self-isolating when required to do so (NHS 2020). One of the key factors determining the effectiveness of such measures is the level of public adherence to them. As the pandemic develops, it is necessary to take into account the impact that a longer duration of measures may have on people’s ability and motivation to continue adhering. It is important also to explore if and how the longer duration of measures may be having on the reasons for non-adherence amongst those who are not fully adhering to them.

There has been much discussion in the media about whether ‘behavioural fatigue’ might be responsible for increases in non-adherence, and whether this will lead to further decreases in adherence in the future (Michie, West & Harvey 2020). However, this concept is controversial in the academic literature, partly because its use has been vague and ill-defined (Harvey 2020) and partly because of the lack of empirical evidence to support the fact that there is significant ‘fatigue’ around adherence. In the UK, longitudinal public surveys generally suggest a lack of evidence that there have been significant decreases in adherence over the course of the pandemic (Michie, West & Harvey 2020; Fancourt et al 2020), or that adherence declined during the summer, before increasing again during the country’s ‘second wave’ (Ipsos MORI 2020). However, these surveys caution that there are high levels of confusion or a lack of understanding of guidelines (Fancourt et al 2020; YouGov 2020). Also, reported ‘complete adherence’ is considerably higher than reported ‘majority adherence’ (where people are following most but not all rules) (Fancourt et al 2020), and some rules (e.g. observing the 1-2 meter rule or visiting extended family when not permitted to do so) may be being flouted more than others (e.g. not self-isolating when advised to do so) (Ipsos MORI 2020).

There is a rapidly growing body of research exploring factors associated with (non-)adherence to coronavirus measures. Surveys of public behaviours suggests that some of the main factors contributing to high adherence include: high perception of risk of contracting coronavirus (Carlucci et al 2020; Kasting et al 2020; Selby et al 2020), perceived social support (Paykani et al 2020), seeing others’ adhere (Coroiu et al 2020), believing that you have already had COVID-19 (Smith, Marteau et al 2020), greater accessing of health-related information (Al-Hasan et al 2020), trust in government (Coroiu et al 2020) and political ideology (Rothgerber et al 2020). Also demographic factors have been found to predict high adherence, including: older age (Selby et al 2020; Fancourt et al 2020), female gender (Carlucci et al 2020), higher education (Carlucci et al 2020; Al-Hasan et al 2020). Of course, there are variations across countries in adherence levels and factors (Al-Hasan et al 2020), due to a variety of reasons, for example, high levels of preexisting trust in authorities and the extent to which a country has a ‘tight’ or collective culture (Van Bavel et al 2020).

Much of this research on adherence to coronavirus measures has taken the form of quantitative surveys. Qualitative research on experiences and perceptions of adherence can complement existing quantitative research by exploring in-depth some of the reasons behind instances of non-adherence, from the perspectives of the participants themselves, which can then be used to adapt policies to maximize impact. In this paper, we present results from an ongoing longitudinal qualitative research study looking at public experiences of, and attitudes to, COVID-19 measures put in place to curb the spread of the virus at a population level. In a previous paper, we explored experiences and perceptions of self-adherence and of adherence in others (Williams et al 2020a). We found that the majority of our participants reported high adherence to measures for themselves, but that reports of non-adherence in others were high (Williams et al 2020a).

The present paper explores two main research questions: (1) What are participants’ perceptions of the current levels of adherence in self and others to COVID-19 measures? (2) What do participants’ feel are the main reasons for non-adherence, in self and others, to COVID-19 measures? Additionally, the paper explores as a secondary aim, participants’ beliefs around levels of future (non-)adherence, as well as the reasons for these beliefs. The study therefore aims to contribute to existing knowledge around current experiences and perceptions of non-adherence to COVID-19 measures.

## PARTICIPANTS AND METHODS

12 focus groups with 51 participants were conducted between 25th September and 13th November 2020. Participants were all adults aged 18 years or older, currently residing in the UK. One of the benefits of conducting focus groups prospectively and in real-time during the pandemic was that it provides data on how participants are feeling in relation to the pandemic, in the moment (thereby avoiding potential recall bias). It is therefore important that participants’ views are qualified and contextualised in relation to rapidly evolving science and policy landscape. Participants’ own views on the changing policy landscape, and its relation to adherence and non-adherence are discussed below. However, by way of context, it is important to briefly note that the study period included a number of important policy developments, including: the introduction of local lockdowns in Wales (introduced on 27 September); Scotland’s 16 day ban on drinking alcohol in licenses premises (introduced on 7th October); the ‘three tier’ system of COVID-19 restrictions in England (introduced on 12th October); the 4 week closure of pubs and restaurants in Northern Ireland (introduced on 14th October); the 19 day ‘firebreak’ lockdown in Wales (introduced 23rd October); Scotland’s five-tier system (introduced 21st October); England’s four-week national lockdown (introduced 5th November). The methodology used in this study has been discussed in previous publications (Williams et al 2020a; Williams et al 2020b). Briefly, online focus groups have both disadvantages and advantages compared to in-person focus groups (Tates et al 2009; Williams 2010), but were obviously necessary in the context of the pandemic, since in-person gatherings were either prohibited by regulation or were generally undesirable in relation to the public health effort (as they were non-essential contact).

Purposive sampling was used to provide as diverse a sample as possible, including country of residence (England, Wales, Scotland and Northern Ireland), gender, age and race and ethnicity (table 1).

**Table 1:** Demographic details reported by participants

| Characteristic | N |
| --- | --- |
| *Gender* |  |
| Female | 25 |
| Male | 26 |
| *Age range* |  |
| 20-29 | 9 |
| 30-39 | 12 |
| 40-49 | 18 |
| 50+ | 5 |
| Not known | 7 |
| *Ethnicity* |  |
| White British | 32 |
| Asian or Asian British | 9 |
| Black or Black British | 4 |
| Other | 6 |

Recruitment took place via a combination of social media advertisements (targeted Facebook ads), online advertising via local community and volunteering sites (e.g. Facebook groups (local community interest groups), Gumtree volunteering groups) and social media snowball recruitment (e.g. via Twitter). Although targeted social media ads attempted to recruit additional members of Black and Asian Minority Ethnic (BAME) participants and those aged over 50, the final sample included a large proportion of white participants aged under 50. Each focus group (average 4 participants per group) met virtually via a web videoconferencing platform (Zoom) for between 60 and 90 min. Participants joined using both video and audio. Focus groups were organised and moderated by SNW and KD. The topic guide for the focus groups was initially developed using existing literature on adherence to health behaviours (discussed above) as well as rapidly emerging research on COVID-19 public attitudes. The main topics for the focus groups included: what people thought about their own adherence and the adherence of others to COVID-19 measures, what they thought the reasons for adherence or non-adherence were, as well as broader topics related to people’s views and experiences around any impact of the pandemic on work life, social life and mental health.

### Analysis

Data were analysed in accordance with a thematic approach as described in Coffey and Atkinson (1996). We took an iterative, pragmatic approach to data collection and analysis, wherein emergent themes from each focus group were used to add to or refine questions during subsequent focus groups. All Zoom focus groups were audio recorded and transcribed. Both authors analysed the transcripts and developed and applied the thematic coding framework. Negative case analysis was used to look for information that did not fit emergent themes, and additional findings and alternative accounts that did not fit with the major themes are discussed in our findings (Silverman 1997). Analysis took a pragmatic approach to grounded theory, whereby initial broad research questions inform the abductive generation of themes (Coffey and Atkinson 1996). Initial primary (open) codes were developed, and were then developed and connected to other related themes to form overarching secondary codes that were developed into the four themes described below (Coffey and Atkinson 1996). Data collection and analysis continued until no new significant themes emerged. Data were analysed in NVivo (V.11.4.3, QSR).

## RESULTS

Most participants felt that, in general, adherence to COVID-19 measures observed in others was currently lower than it has been earlier in the pandemic. In particular, participants reported seeing others failing to stay 1-2 metres apart (especially in shops) and knowing of others meeting with family and friends in households (in ways not permitted at the time). As was observed earlier in the pandemic (Williams et al 2020a, reports of non-adherence in self were lower compared to reports of non-adherence in others. However, reports of non-adherence in self were higher at this stage of the pandemic as compared to reports of non-adherence in self at an early stage of the pandemic (cf. Williams 2020a). In terms of their own adherence, most participants described trying to adhere to measures as best they could, but discussed a number of ways in which following them had been challenging. Additionally, most participants felt that adherence in others was likely to decline further in the near future, particularly around the Christmas holidays, with some suggesting that self-adherence would also be lower. Analysis revealed six main themes related to participants’ reported reasons for non-adherence, where it was being observed: (1) *Alert fatigue* (2) *Inconsistent rules* (3) *Lack of trust in government* (4) *Helplessness* (5) *Resistance and rebelliousness (6) Reduced perception of risk and the prospect of a vaccine*. No obvious patterns emerged according to the demographics of the participants, with a mix of genders, ages and races and ethnicities being represented in each theme. In this section we describe these themes, before further exploring their interrelations and implications in the discussion section.

### Alert fatigue

One of the main reasons for non-adherence, particularly in relation to non-adherence in self, was the high volume and frequency of information that they had been exposed to over the course of the pandemic. Specifically, the frequent government announcements related to what was often perceived to be constantly changing and complex rules had left many feeling “lost” (Participant 7, Female, 20s) or “confused” (Participant 31, Male, 40s), and that it was “impossible to keep up with the rules” (Participant 29, Female, 40s). As one participant put it, “I’m feeling a bit fatigued, sick of it” (Participant 13, Male, 30s). This constant stream of information could be seen to be causing information fatigue, or rather a particular form of information fatigue referred to as *alert fatigue* (Cash 2009). This concept is derived from research on clinical decision making support systems, and its understood as the mental state that comes from receiving too many alerts that consume time and energy which can cause important alerts to be ignored along with clinically unimportant ones (Peterson and Bates 2001; Phansalker et al 2012). In our case, alert fatigue was observed where participants felt it was difficult to follow (“keep up with”) or remember what the rules were. This in turn meant that potentially important information was being missed or not taken as seriously as it might or should have:

> *“It’s been hard this lockdown, with all these government measures and restrictions. … I don’t know what the new legislation is. It’s hard to keep up-to-date with all of this information. I’ve come to the decision where I am going through day-by-day, but placing less attention on whatever the government is saying…. I’m not as concerned about following the measures as seriously as I should have done*.*” (Participant 2, Male, 30s)*

Whereas government announcements (“conferences”) had once been given significant attention and been taken seriously by the public, the cumulative effect of the over-exposure to these alerts was leading participants to become desensitised or habituated to this information. Participants felt that the constant exposure to new information on the pandemic, meant that it became hard to distinguish between important announcements about new rules that directly affected them, and less important or superfluous information (“just updating”):

> *“It does feel like they are changing the rules like every week, and then people don’t take them seriously, and I don’t blame them…. There have been too many conferences … I go on Facebook and it’s coming up every few days as a notification ‘Boris [Johnson, UK Prime Minister] is live’ … I never watch them anymore because I don’t know if they are going to be new rules or whether he is just updating us, I don’t know what’s going on … If we just had fewer and they concentrated just on the rules, individuals might concentrate more and take it more seriously*.*” (Participant 8, Female, 20s)*

The sense of information fatigue over the frequency of announcements and rule changes was compounded for some participants by the fact that information wasn’t being communicated clearly enough, partly because of the overload of information *per se*, and partly because much of it was technical and esoteric information that was insufficiently explained or translated:

> *“I watch the [Welsh Government] announcements on Facebook Live stream, and after he [Mark Drakeford, Welsh First Minister] talks, it just seems like a Q and A for an hour, with all these long words, and you haven’t got any further with what the rules are, and you are left feeling ‘what does that mean?’” (Participant 39, Female, 20s)*

### Inconsistent rules

In addition to feeling over-exposed to information about COVID-19 measures, participants also expressed feeling as though measures were often “confusing because of the mixed messages” (Participant 3, Male, 20s). This was discussed both in relation to non-adherence in self and in others. One of the main causes of this confusion was the perception that rules were inconsistent, either because they were changing so much over time (as discussed above as a cause of alert fatigue) or because they were inconsistent across place (i.e. between countries in the UK and between different regions within each country). Participants criticised what they saw as the inability, or unwillingness, of political leaders to create consistent policy and present a unified front:

> *“I’m most upset with our individual home country leaders. The fact that they cannot get on the same page, the rules are already confusing, but then they make them more confusing … they should have been able to come up with one set of rules, it’s absolutely insane” (Participant 13, Male, 30s)*.

Participants also expressed concern that the lack of consistency would also be a problem for the future:

> *“I don’t feel like anybody is taking anything very seriously, because one rule is being made and another one contradicts it. … I couldn’t care less… People aren’t really aware of what they are doing or why they are doing it*.. *especially with these new rules about the tiers, I’ve just not bothered to even look into them, because they are not making sense to me. Everywhere has adopted the rules, but then they have refined them, changed them and interpreted them to suit themselves. Some of them make sense and some of them don’t. … We don’t know the rules for Christmas. And so, is that our life now? That we have to wait for different rules, like every month, rules that apply to some and rules that don’t apply to others, and is this how we are going to live for the next few years?” (Participant 28, 50+)*

Non-adherence in this sense was not constructed as non-conformity, insofar as people were overtly and deliberately breaking rules, but rather as a process of s*ubjective rule interpretation*. This was seen to be outcome of the lack of clarity and consistency in the way in which rules were made and communicated. As above quote suggests, some participants felt that people were (perhaps intentionally) exploiting the lack of clarity and consistency. Others however felt that subjective rule interpretation was an unintentional, and perhaps unavoidable, consequence of the inability to make sense of, or keep up with, the rules:

> *“I’m not some total non-conformist. Yes, the rules are there to help, and I try to abide by them, but it’s just gone to the extreme now where, if I asked the three of you now what the rules are, we would probably all say something different*.*” (Participant 6, Male, 20s)*

### Lack of trust in government

Participants also discussed their own, and others’, relative lack of adherence to measures in relation to a general lack of trust in, or “respect” for, the UK government, who were seen to have “handled the pandemic very badly” (Participant 29, Female, 40s). The lack of trust in government was seen as something that encouraged the more intentional exploitation of the rules, as discussed above. It was felt that a lack of respect in authority permitted people to subjectively interpret (“make up”) their own rules, perhaps to fit, or justify, their desires to engage in behaviours that were not permitted within the rules:

> *“We don’t respect our government, we don’t respect the decisions they are making, we don’t respect their rules. We might follow something that we decide to follow, but then once we are allowed to say ‘oh yeah but its Christmas’ or ‘oh but somebody’s getting married’ and ‘we need to do this, we need to do that’, then everybody makes up their own rules and goes ‘well we are going to get away with it anyway. … If figures [case rates] are going up, and if they continue to go up, by Christmas, I am not sure if any rule they make is going to be appropriate*.*” (Participant 28, 50+)*

In this way, the lack of respect for government, combined with the belief that rules were inconsistent or ambiguous was a means of reconciling the cognitive dissonance (the mental conflict arising from a mismatch of belief and behaviour) (Festinger 1957) that others were assumed to be feeling. That is, participants often reported feeling torn - between wanting to socially integrate but also knowing that doing so might not be in the interests of public health. As the above quote illustrates, rules were often seen to contain exceptions or loopholes that could be exploited (i.e. “make up their own rules”), something that negatively impacted motivation to adhere or permitted people to subjectively interpret the rules to permit or justify desired behaviour (e.g. meeting more people indoors than might be permitted at Christmas).

A key reason for the loss of trust was the well-publicised instances in which politicians were seen to be subjectively interpreting the rules to their own benefit. Participants argued that if those in positions of authority were unable or unwilling to follow rules, why should the public be expected to do so. This lack of trust, it was argued, was exacerbating the stress people were experiencing during the pandemic:

> *“There is no doubt that the way the government is communicating and handling things is going to add to people’s stress. It’s inevitable, it’s happening now … it doesn’t help that the politicians are interpreting the rules to their own benefit as well*.*” (Participant 31, Male, 40s)*

### Helplessness

Another major theme related to non-adherence in both self and others was a feeling of helplessness (being “fed up” and “giving up”). This feeling of helplessness was in some respects a product of, or response to, the alert fatigue, inconsistent rules, and lack of trust in government discussed above. For example, some linked their feeling of helplessness, and the associated emotional toll (feeling “down”) to the over-exposure to news on COVID-19, including the frequently changing rules, and the frequent news coverage them, discussed above (alert fatigue):

> *“I have just given up. I don’t even look at the news, I mean it changes everyday and I’m fed up. It just gets you down really” (Participant 29, Female 40s)*

Some participants reported “struggling to have a positive outlook on life” (Participant 6, Male, 20s) and generally feeling pessimistic over the future, particularly in regard to how long they would be subject to some form of coronavirus measures. This included looking ahead to Christmas and the new year, which they characterised as being normally happy times, but which would be different this year:

> *“We are going into the New Year, and normally we have that feeling of setting new goals, and having new things to look forward to. What is the mental state of people going to be? We are going to enter 2021 and people are going to have so much uncertainty about the new year and covid*.*” (Participant 25, Male, 40s)*

Participants tended to attribute people’s sense of helplessness to a loss of control over their lives:

> *“People have lost control over their life*… *and when you lose control over your life, thats it*.*” (Participant 15, Male 40s)*

For some, the control had been taken from them by government regulation. They expressed a sense of resentment for the way in which, as they perceived it, general civil liberties were being “taken away” (for example, the freedom to socially interact in ways of their choosing), at the same time as “small” freedoms were being “given back” (for example, related to the debate over whether at Christmas people might be permitted more freedom):

> *“I am not happy that they take things away from you and then find little ways of giving bits of it back. Like you should be so grateful that you are allowed out for Christmas, but not things like that; it’s getting very weird, and I wish they would be more truthful and just said, don’t mix with anybody, use your common sense*.*” (Participant 28, 50+)*

For a number of participants, the loss of autonomy and control over their lives (“they take things away from you”), compounded by the inconsistency (“giving bits of it back”) and the lack of trust in government (“I wish they would be more truthful”), led them to follow (or advocate for) “common sense”, as opposed to official rules. This feeling of helplessness, of a lack of control or optimism for the future, had been accumulating over the course of the pandemic. One of the main factors for this was the constant uncertainty surrounding whether measures would change, and ultimately how long some combination of measures would be in place. The feeling of there being no defined end point (“is this our life now?”) added to the sense of helplessness. Participants linked this uncertainty and the growing feeling of helplessness to observations of non-adherence, of “giving up” trying to strictly follow the rules (“we might as well do what we want”):

> *“I think at the beginning [of the pandemic], we were told it was three weeks, which then turned to three months, which now has turned to six months, and this is so past what anyone could have expected, everyone has thrown in the towel, like ‘we don’t know when it’s going to end, we are going to give up on waiting, because we have been waiting so long, so we might as well do what we want without getting fired or arrested … and I think now that we are talking about Christmas and it’s not going to happen … and so I think people have lost hope and just half accepted how this is and are going back to a normal life as best they can instead of being really strict and taking it seriously*.*” (Participant 8, Female, 20s)*

### Resistance and rebelliousness

Another theme that emerged as a reason for non-adherence, particularly non-adherence in others, was people’s resistance and rebelliousness in relation to COVID-19 measures. Often this was seen to take the form of *overt rule-breaking*. Participants viewed it as a consequence of, or response to, the feeling of being “fed up” or a loss of motivation to continue to adhere:

> *“I’m hearing stories of people being rebellious…. I think in general there is a general feeling of being fed up… there is a lady here who would be outspoken about the rules, if people were breaking them, but even she was last night saying she was away with her daughter so now even she is breaking them*.*” (Participant 26, Female, 40s)*

Participants felt this would worsen in the future, particularly around the Christmas holidays:

> *“I think a lot of people won’t [stick to the rules], I think they will say ‘you know what I want to enjoy my Christmas and they are going to break the rules, even if it is slightly. I think if you have got a family, and people are told you can’t mix with another household unless it is outside, people are going to say ‘Its Christmas and I want to see my family”. People are going to say I don’t care, I want to have something to look forward to*.*” (Participant 30, Male 40s)*

For some, this overt rule-breaking was a form of protest to the loss of autonomy and control (“we don’t like following rules”) and the inconsistent rules (“the mixed messages”), and was something that would worsen in the future, potentially even taking the form of “civil unrest” or “protest” (Participant 49, 50+):

> *“And really it is getting on for nearly a year come Christmas time. I mean how long can you go without seeing people? I’ll go to visit [family’s town]. People are going to break the rules, of course they are. It is going to get even worse, where people are going to get completely fed up, so I can’t really see anybody following the rules in December to be honest …. We are quite bolshy in the UK. We like a riot and don’t like following rules … there is going to be civil unrest, because there is already in other countries, and it’s because of the mixed messages*.*” (Participant 29, Female 40s)*

For some, resistance was the attempt to re-exert control over their lives, to reassert their autonomy, by focusing on controlling what they could control. In some instances, this included overt rule-breaking (e.g. not wearing masks when required):

> *“The more you know you can’t really change the outcome of it, all you can do is make the most of your own life and adapting to it. I feel very overwhelmed … I don’t want to talk to people because I don’t want to get into an argument about it … I’m not wearing a mask and I’m not going to. I’m going to stick to what I believe … I am responsible for my life and my daughter*.*” (Participant 26, Female, 40s)*

Others felt that resistance would take form of subjective rule interpretation, rather than overt rule-breaking. They also however framed non-adherence as a response to the loss of autonomy and control, as something that would worsen in the future (also potentially taking the form of “civil unrest”):

> *“I can see this getting worse with Christmas approaching, because people, fair enough, will not stand for being controlled. People will want to travel and see their loved ones … If we are in a tiered system people will try and interpret the rules in their own way … we are not great at being dictated to, or doing things that are for our benefit… and it’s going to get worse from a civil unrest point of view. I fear we are sleepwalking into a police state*.*” (Participant 31, Male, 40s)*

Many participants discussed examples of ways in which people were engaging in small, creative and subtle forms of rebellion to government rules. This included engaging in social rituals now perceived as deviant (e.g. handshaking, not wearing masks when required to), or mis-appropriating new rituals (eg. improperly wearing masks) as subtle forms of passive resistance to rules:

> *“You have got some people who are like ‘covid is here’, you are seeing the masks, but then you have got this other spectrum where it is almost like a rebellion against covid, where people are like, and I kind of understand it. Like with local lockdowns where you can’t go there but you can go here, you have gone from being told you don’t need to wear a mask because they don’t make any difference, to you must wear a mask everywhere you go… I had a guy at the burger van the other day, and he goes ‘alright [Name], how are you?’ And he goes to shake my hand’. And I said ‘er, social distancing?’ And he goes ‘I don’t believe in that shit’ and he just grabbed my hand and shook it*,. *And I was like ‘what are you doing, now I’m going to have to wash my hand!’” (Participant 41, Male, 30s)*

### Reduced perception of risk: The prospect of a vaccine

As discussed in the methods section above, one benefit of conducting qualitative research in real-time during the pandemic is the opportunity to capture emerging themes in a rapidly evolving policy and scientific landscape. One example of this was the announcement on the 9th November 2020, that a leading vaccine candidate (developed by the pharmaceutical and research companies *Pfizer* and *BioNTech*) had announced Phase 3 clinical trial results which indicated that their COVID-19 vaccine was 90% effective in preventing the disease, and might start being rolled out as early as the end of the year (The Guardian 2020). In the five focus groups conducted after the announcement, an additional theme which emerged was the participants’ perceptions of how the prospect of a vaccine might serve to reduce the perceived risk, or threat, of COVID-19, and how this might adversely impact public adherence to measures. Views coded under this theme tended to focus on the fact that the announcement made the prospect of a vaccine more tangible, and that the pandemic was losing its “fear factor” (Participant 13, Male 30s). This theme was nearly always focused on adherence in others (with no participants reporting that it would make them less likely to adhere). Some participants felt this would have a general impact on adherence:

> *“I get the impression people are going to be more relaxed, like ‘oh there is a vaccine now, the problem is fixed. I went to Tesco twice today and both times I saw people without masks for the first time [since it was made compulsory]. I personally think it [the vaccine] takes the danger away*.*” (Participant 38, Male, 30s) “I think people will be more blasé now, or people will think ‘we can carry on now because we have a vaccine, we will be fine*.*’” (Participant 46, 50+)*

This could be compared to arguments earlier in the pandemic around mask-wearing, where concerns existed that mask wearing might encourage a false sense of security thereby discouraging people from maintaining good hygiene or social distancing behaviours (Lazzarino et al 2020). Other participants felt that news would only impact the adherence of those who were already prone to non-adherence (i.e. it would serve to justify and increase their non-adherence):

> *“I think of the people who are currently feeling that way inclined, you know to bend the rules, break the rules, stretch the rules, they probably will break the rules more. However, I wouldn’t say that about everybody. There are groups who are still taking things very seriously, still observing the guidance and just because the vaccine has been announced I don’t think people are going to go and stop. I personally feel like we don’t have enough information to know if it is going to be the golden bullet … But those who are inclined to break the rules are more likely to take the news of a vaccine to mean ‘it’s all over lets crack on with life*.*’” (Participant 34, Female, 30s)*.

### Alternative accounts on (non-)adherence

Although the themes discussed above constituted the prevailing views expressed during the focus groups, negative case analysis suggested that some participants held different related to adherence and non-adherence. For example, although most participants argued that adherence in general was waning over time, some participants offered alternative accounts. Some participants for example argued that adherence to social distancing had been high during the first lockdown, had lessened during the summer, but was now being “taken more seriously again” (Participant 40, 50+). They attributed this to various factors, including most notably the perception of risk had fallen and then risen again corresponding to the number of cases between waves. Some participants described how for them personally, adherence hadn’t changed much over the course of the pandemic. These participants tended to characterise themselves as being quite strict or conscientious about the rules and as such felt that they hadn’t changed their behaviour (e.g. “we never went fully out, we never went to a restaurant” (Participant 11, Female 40s) during the summer when measures had relaxed. Also, not all participants felt that policy had been inconsistent, hard to understand or follow. Fr example, some participants “liked” the tier system because it helped them better “understand what risk we are at in relation to those around us” (Participant 10, Male, 30s).

## DISCUSSION

Six themes emerged from our grounded analysis across all focus groups: Alert fatigue; inconsistent rules; a lack of trust in government; helplessness; resistance and rebelliousness; and (in later focus groups) reduced perception of risk due to the prospect of a vaccine. These themes are not mutually exclusive and are inter-related in complex ways. For example, in some instances the feeling of helplessness was attributed to the frustration of not being able to “keep up” with rules (alert fatigue) or the feeling they were inconsistent or didn’t “make sense”. Also, the resistance and rebelliousness reported was in part seen as a response to the frustration over inconsistent policy and a lack of trust in or respect for government, compounded by the perceived loss of autonomy and control. Taken together these factors were associated with reduced adherence to rules either in the *form of overt rule-breaking* or *subjective rule-interpretation*.

Recent survey evidence suggests that self-reported majority adherence in the UK has generally remained high and stable throughout the pandemic (Fancourt 2020a). It may be the case that discrepancies exist between (high) adherence in self and (low) adherence in others (cf. Williams 2020a). A number of factors could account for this discrepancy, including social desirability bias (the desire to appear conscientious) or selection bias (those more likely to adhere might also be more likely to take part in a research study on adherence).

However, our findings suggest that, from the perspective of our participants, non-adherence in self and others may be more common as compared to earlier stages in the pandemic (cf. Williams 2020a). All six themes were observed in related to both non-adherence in self and non-adherence in others. However, certain themes were more prominent in relation to participants’ accounts of non-adherence in self as compared to non-adherence in others (and vice-versa). For example, participants were more likely to discuss their own non-adherence in terms of alert fatigue or subjective rule interpretation and others non-adherence in terms of resistance and rebelliousness and overt rule-breaking. Perhaps one explanation for this is the fundamental attribution error, or specifically the actor-observer bias (Watson 1982) which the holds that people have a tendency to believe others’ actions stem from stable characteristics (e.g. rebelliousness) while seeing their own actions to also stem fro contextual factors (e.g. an overload of complex information). It is important to note however that inconsistent policy and a lack of trust was pervasive in both self and others’ non-adherence.

Although, as discussed above, the notion of ‘behavioural fatigue’ is a largely vague and poorly evidenced concept (Michie, West & Harvey 2020), our findings do suggest that a very specific form of fatigue - *alert fatigue* - may be an important factor for non-adherence to guidelines (amongst those who are not adhering). This phenomenon is variously explained as a result of cognitive overload (where too much information cannot be processed, or retained, effectively) or desensitisation (where repeated exposure to alerts leads to declining responsiveness to them) (Anker et al 2017). In their study of public health care providers, Baseman et al (2013) argued that “during a pandemic when numerous messaged are sent, alert fatigue may impact ability to recall when a specific message has been received due to the ‘noise’ created by the higher number of messages”. Our findings suggest that in the context of the current coronavirus pandemic, alert fatigue is a wider phenomenon being experienced by some within the general public. Indeed, the concept of alert fatigue has its roots in the much broader concept of “information overload” which was originally taken to be where a person develops a “blasé outlook” as a means to cope with a constant influx of stimuli (Simmel 1903). Many of our participants described how they were no longer paying as much attention to - or even actively avoiding - to what they perceived as “constant” news on COVID-19, including what was perceived as excessively frequent and detailed announcements (alerts) made by political leaders. As such, in their desire to provide “transparency”, and to communicate “the science” behind policy, government may have counter-productively “overloaded” the public with information to the point at which some are actively avoiding it. Such avoidance could be construed as a coping mechanism both in regard to the cognitive overload (of having to filter through the “noise”) and in regard to the anxiety or stress caused by the focus on the pandemic and the measures.

Of course, the frequency of the alerts was also related to the fact that policies were also changing frequently. Our findings suggest that participants were often confused over what they perceived to be constantly changing rules, which were sometimes inconsistent or didn’t “make sense” to them. Survey evidence is beginning to suggest that more than half (55%) of the UK public have found the rules to be confusing (YouGov 2020). Our findings have explored qualitatively some of reasons why participants have found the rules confusing. The differing policy approaches and timelines between the four nations was a key source of confusion, as was the introduction of local lockdowns and, in England, the tier system. Significantly, the geographic variability, in addition to the variability over time, was perceived by some as being a reason as to why they were no longer able to, or no longer chose to, follow “the rules”.

Related to both alert fatigue and inconsistency in rules was a general sense of a lack of trust in, or respect for, government. The consistent decline in confidence in the UK government has been documented in longitudinal surveys (YouGov 2020; Ipsos MORI 2020). Additionally, it is well-established that role modelling by people in positions of leadership is an important factor in motivating adherence, and research on COVID-19 is starting to explore the adverse impact of poor behavioural role modelling (the so-called “Cummings Effect” (Fancourt et al 2020)). As seminal research in psychology has shown, trust and confidence in those in authority plays a key role in the extent to which they are followed (Milgram 1971), as does positive role modelling (Bandura 1986). Milgram (1971) found that polyphony - that is the disagreement between conflicting voices of authority - negatively impacts compliance by increasing the likelihood that people will have to decide for themselves what action to take. In the case of COVID-19 measures, there has been an increasing trend towards polyphony amongst political leaders across the UK, something participants noted when contrasting recent measures to the initial UK-wide lockdown in March. The existence of frequent announcements about different countries and regions from multiple voices (e.g. national and regional political leaders) likely adds to the alert fatigue some may be experiencing. Our findings suggest that, from the perspective of our participants, a more unified set of measures across the UK, or at least the appearance of a more collaborative and unified voice, would have resulted in an improved capacity to understand, and thus potentially follow, rules.

The theme of helplessness was also prominent in our findings. Although our previous research, conducted during the first wave of the pandemic explored how social distancing was already leading to feelings of ‘loss’ which in turn was having negative impacts on mental health, feelings of helplessness and a loss of control did figure prominently at the time (Williams et al 2020a). This helplessness appears to have emerged as a result of the *sustained* social sacrifice and social suffering (a sustained sense of loss), coupled with a growing sense of a lack of control, as the pandemic has drawn on. As such, it might be understood in relation to Seligman’s concept of learned helplessness (Abramson, Seligman & Teasdale 1978), insofar as some participants were being ‘conditioned’ to feel as though they had little control over the outcome of the pandemic and over their lives in general (drawing a comparison to seminal experimental studies (Maier & Seligman 2016), they had been experiencing a ‘failure to escape’ social restrictions loss of freedoms). This failure to escape and loss of control has been linked to the symptoms of depression such as hypersomnia, hyperphagia, excessive guilt (attempt to personalize control) and low self-esteem (I should be doing better). Similar to ‘giving up’ in the context of health behaviours, such as giving up trying to stop smoking, helplessness in the context of covid measures meant ‘giving up’ trying to adhere to the rules. Such helplessness stems from cognitive, affective and motivational deficits (Strecher et al 1986) which some of our participants reported in the form of a perceived lack of ability to understand, or motivation to adhere to, rules. Perhaps even more aptly, this helplessness can be understood in relation to Bandura’s (1986) social cognitive theory. Over the course of the pandemic, outcome expectations over the hoped-for success of social distancing restrictions were not being met (“this is so past what anyone could have expected”). At the same time, perceptions of self-efficacy to sustain adherence were being adjusted (“we are going to give up on waiting … we might as well do what we want”).

Our findings suggest that resistance and rebelliousness may be becoming more observable – at least in regard to people’s perceptions of non-adherence in others - as resilience to coronavirus measures is increasingly tested. Indeed, resistance can be seen as a natural corollary of control (Foucault 1975), particularly where such control is felt to be excessive or sustained. Our findings reveal the ways in which some people may be performing ‘resistance through rituals’ (Hall 1975), by for example eschewing or mis- appropriating new rituals (e.g. failing to wear masks or wearing them improperly) or by intentionally engaging in rituals that were once normative but are now increasingly considered deviant (e.g. handshaking or not keeping physically distant). This rebelliousness might be characterised as ‘mis-behaviour’ that ‘arises from an impulse to take control rather than to be always subject of control … and seldom comes simply from a desire to break rules’ per se (Ackroyd & Thompson 1999

Finally, our results suggest that the prospect of a vaccine might have an adverse impact on adherence, either by lowering the overall perception of risk in the general public, or by encouraging those who were already prone to non-adherence to continue to do so. Research evidence, on COVID-19 as well as in east outbreaks of infectious disease, e.g. pandemic influenza, suggests that one of the key drivers. The prospect of a vaccine in the near future could serve to reduce the perceived risk of the virus. This theme, as with the others identified in our study, warrant further investigation in future research.

### Limitations

One limitation of this study is that it has likely overlooked a range of other factors that relate to (non-)adherence. As a qualitative and grounded analysis, we discuss only those most prominent themes that emerged from our particular data set. As discussed above, the nascent literature on COVID-19 policy adherence has found a range of other factors which have not been identified or discussed in the present study (e.g. Carlucci et al 2020; Kasting et al 2020; Selby et al 2020). Also, although this study did not identify any patterns by demographic variables, this is potentially a result of relatively small sample size of a qualitative study with a diverse group of participants. It does not challenge the notion that life circumstances - for example, an individual’s socio-economic status, age, geographic location (etc) - play an important role in adherence, and patterns reflecting this may be best identified by large sample quantitative surveys (e.g. Fancourt et al 2020). Another limitation of this study is that did not recruit participants from clinically extremely vulnerable or clinically vulnerable categories, for example, individuals aged 70 and over and those living with those with particular serious health conditions. Also, although our recruitment material did encourage those at high risk to apply, we received no applications from those over 70. This may be due to the fact that these are a hard-to-reach group online who are significantly less likely to use social media or be heavy internet users (van Boekel et al 2017).

### Implications for policy and practice

Our study has a number of implications for understanding non-adherence to COVID-19 measures in instances where it is occurring. Firstly, participants felt that news of an effective vaccine might lead some people to be more complacent about social distancing, even before the vaccine is widely available, because of the loss of the virus’ “fear factor”. Secondly, some participants were prepared to break rules over Christmas if they feel they are too restrictive and particularly if they prevent family gatherings. Also, many participants felt that rule-breaking over Christmas amongst the general public will be higher than it is currently. Thirdly, a number of participants reported feeling alert fatigue after experiencing months of rule changes, announcements and news on COVID-19 and are not following or understanding official government rules as a result. Fourthly, many participants found that official government rules were too inconsistent and complicated. Fifthly, participants reported a lack of trust in or respect for government. For some, these factors may be leading of a sense of “learned helplessness”, where they feel like they have “given up” keeping up with and following covid rules. For others, they may be leading to increasing feelings of resistance and rebelliousness, with some concerned that sustained lockdown measures would increasingly bring about “civil unrest”.

Finally, the study sheds light on two forms of non-adherence around COVID-19 rules. Although some non-adherence is likely taking the form of overt rule-breaking (perhaps largely as a form of resistance to the loss of control), much of the non-adherence observed in and by our participants takes the form of subjective rule interpretation. In order to improve adherence, governments, health officials should as far as possible seek to: (1) Strike a balance between open and transparent communication around measures and ‘overloading’ the public with information around rules and rule changes; (2) implement a more unified set of measures across the UK and a consistent message (voice) from political leaders (taking into consideration the potential confusion related to the tier system for some); (3) work to rebuild public trust, through exemplary adherence to rules amongst those in positions of authority; (4) develop measures that take into consideration that motivation to adhere to very restrictive rules might be lower during the Christmas period; and (5) ensure that public health measures and messages take into consideration the fact that news of vaccine ‘breakthroughs’ may negatively impact some people’s adherence to current and future measures.

## Data Availability

Ethical restrictions related to participant confidentiality prohibit the authors from making the data set publicly available. During the consent process, participants were explicitly guaranteed that the data would only be seen my members of the study team. For any discussions about the data set please contact the corresponding author, Simon Williams.

## DECLARATIONS

### Transparency declaration

The lead author (the manuscript’s guarantor) affirms that the manuscript is an honest, accurate, and transparent account of the study being reported; that no important aspects of the study have been omitted; and that any discrepancies from the study as planned (and, if relevant, registered) have been explained.

### Authors’ contributions

All authors contributed to the planning of the study. The analysis was conducted by SW and KD. The initial draft of the article was written by SW. All authors revised the manuscript and approved the final version for publication. SW is the guarantor of the article.

### Data sharing statement

Ethical restrictions related to participant confidentiality prohibit the authors from making the data set publicly available. During the consent process, participants were explicitly guaranteed that the data would only be seen my members of the study team.

For any discussions about the data set please contact the corresponding author, Simon Williams.

### Ethics statement

Ethical approval was received by Swansea University’s School of Management Research Ethics Committee.

## REFERENCES

Abramson, L. Y., Seligman, M. E., & Teasdale, J. D. Learned helplessness in humans: Critique and reformulation. Journal of Abnormal Psychology 1978;87(1), 49–74.

Ackroyd S., Thompson P. Organizational Misbehaviour, London, Sage, 1999.

Al-Hasan A, Yim D, Khuntia J. Citizens’ Adherence to COVID-19 Mitigation Recommendations by the Government: A 3-Country Comparative Evaluation Using Web-Based Cross-Sectional Survey Data. J Med Internet Res 2020;22(8):e20634

Ancker, J.S., Edwards, A., Nosal, S. et al. Effects of workload, work complexity, and repeated alerts on alert fatigue in a clinical decision support system. BMC Med Inform Decis Mak 2017;17, 36.

Bandura A. Social Foundations of Thought and Action: A Social Cognitive Theory. Englewood, NJ, PrenticeHall, 1986.

Baseman, J.G., Revere, D., Painter, I. et al. Public health communications and alert fatigue. BMC Health Serv Res 2013;13, 295.

Carlucci L., D’Ambrosio, I., Balsamo, M. Demographic and Attitudinal Factors of Adherence to Quarantine Guidelines During COVID-19: The Italian Model. Frontiers in Psychology 2020;2(11), 2702.

Cash J. Alert Fatigue. American Journal of Health-System Pharmacy,66, Issue 23, 1 December 2009, 2098–2101.

Coffey A, Atkinson P. Making Sense of Qualitative Data. London: Sage, 1996.

Coroiu A, Moran C, Campbell T, Geller AC. Barriers and facilitators of adherence to social distancing recommendations during COVID-19 among a large international sample of adults. PLoS ONE 2020;15(10), e0239795.

Fancourt D, Bu F, Mak H, Steptoe A. UCL COVID-19 Social Study Results Release 22 [Internet]. 2020a Available from: https://b6bdcb03-332c-4ff9-8b9d-28f9c957493a.filesusr.com/ugd/3d9db5_636933e8191d4783866c474fab3ca23c.pdf

Fancourt D, Steptoe A, Wright L. The Cummings effect: politics, trust, and behaviours during the COVID-19 pandemic. Lancet. 2020b;396(10249):464–5.

Festinger, L. A theory of cognitive dissonance. Stanford, C.A., Stanford University Press. 1957. Foucault M. Volume I: The History of Sexuality. New York, Pantheon Books, 1978.

Gonzalez V., Goeppinger J., Lorig, K. Four Psychosocial Theories and Their Application to Patient Education and Clinical Practice. Arthritis Care and Research 1990; 3(3), 132–143.

Harvey N. Behavioural fatigue: real phenomenon, naive construct or policy contrivance. Frontiers in Psychology. 2020. Available at: https://www.frontiersin.org/articles/10.3389/fpsyg.2020.589892/abstract

Hall S., Jefferson,T. (eds). Resistance Through Rituals Youth subcultures in post-war Britain. London, Routledge, 1975.

Ipsos MORI. Britons increasingly abiding by the COVID-19 rules, with social responsibility and the NHS the primary drivers. 2020. Available from: https://www.ipsos.com/ipsos-mori/en-uk/britons-increasingly-abiding-covid-19-rules-social-responsibility-and-nhs-primary-drivers

Kasting ML, Head KJ, Hartsock JA, Sturm L, Zimet GD. Public perceptions of the effectiveness of recommended non-pharmaceutical intervention behaviors to mitigate the spread of SARS-CoV-2. PLoS ONE 2020;15(11): e0241662.

Lazzarino AI, Steptoe A, Hamer M, Michie S. Covid-19: important potential side effects of wearing face masks that we should bear in mind. BMJ 2020;369(m1435):m2003. doi:10.1136/bmj.m2003 pmid:32439689

Maier S., Seligman M. Learned Helplessness at Fifty: Insights from Neuroscience. Psychol Rev. 2016;123(4), 349–367.

Mantzari E., Rubin GJ, Marteau T. Is risk compensation threatening public health in the covid-19 pandemic? BMJ 2020;370:m2913.

Michie S., West R., Harvey N. The concept of “fatigue” in tackling covid-19. BMJ 2020;371:m4171

Milgram, S. Obedience to authority: An experimental view. New York. NY: Harper & Row, 1974.

National Health Service (NHS) UK. Guidance on social distancing. https://www.nhs.uk/conditions/coronavirus-covid-19/social-distancing/what-you-need-to-do/ [Accessed 12 November 2020].

Paykani, T., Zimet, G.D., Esmaeili, R. et al. Perceived social support and compliance with stay-at-home orders during the COVID-19 outbreak: evidence from Iran. BMC Public Health 2020;20, 1650.

Peterson JF Bates DW. Preventable medication errors: identifying and eliminating serious drug interactions. J Am Pharm Assoc 2001;41:159–60.

Phansalkar S., et al. Drug—drug interactions that should be non-interruptive in order to reduce alert fatigue in electronic health records. J. Am. Med. Inform. Assoc. 2013;20:489–493.

Rothgerber, H., Wilson, T., Whaley, D., Rosenfeld, D. L., Humphrey, M., Moore, A. L., & Bihl, A. Politicizing the COVID-19 Pandemic: Ideological Differences in Adherence to Social Distancing. Psyarxiv 2020. https://doi.org/10.31234/osf.io/k23cv

Selby K, Durand MA, Gouveia A, Bosisio F, Barazzetti G, et al. Citizen responses to government restrictions in the COVID-19 pandemic: a cross-sectional survey in Switzerland. JMIR Form Res. 2020 doi: 10.2196/20871. Epub ahead of print.

Silverman D. Qualitative research: theory, method and practice. London: Sage, 1997

Simmel G. The Metropolis and Mental Life. adapted by D. Weinstein from Kurt Wolff (Trans.) The Sociology of Georg Simmel. NewYork: Free Press, 1950, pp.409–424.

Strecher VJ, Devellis, BM, Becker, MH, and Rosenstock, IM: “The Role of SelfEfficacy in Achieving Health Behavior Change.” Health Education Quarterly, 1986;13(1),73–92.

Tates K, Zwaanswijk M, Otten R, et al. Online focus groups as a tool to collect data in hard-to-include populations: examples from paediatric oncology. BMC Med Res Methodol 2009;9:ar15.

van Bavel, J.J.V., Baicker, K., Boggio, P.S. et al. Using social and behavioural science to support COVID-19 pandemic response. Nat Hum Behav 2020;4, 460–471.

van Boekel LC, Peek ST, Luijkx KG. Diversity in older adults’ use of the Internet: identifying subgroups through latent class analysis. J Med Internet Res 2017;19:e180.

Watson, D. (1982). The actor and the observer: How are their perceptions of causality divergent? Psychological Bulletin, 92(3), 682–700.

Williams SN. A twenty-first century Citizens’ POLIS: introducing a democratic experiment in electronic citizen participation in science and technology decision-making. Public Underst Sci 2010;19:528–44. 14

Williams SN, Armitage CJ, Tampe T, Dienes KD. Public perceptions and experiences of social distancing and social isolation during the COVID-19 pandemic: a UK-based focus group study BMJ Open 2020a;10:e039334.

Williams, SN, Armitage, CJ, Tampe, T, Dienes KD. Public attitudes towards COVID-19 contact tracing apps: A UK-based focus group study. Health Expectations 2020b; forthcoming. YouGov. COVID-19 Public Monitor, Available at: https://yougov.co.uk/covid-19 (Accessed 20 October 2020)

